# Predicting Response to Radiotherapy of Intracranial Metastases with Hyperpolarized ^13^C MRI

**DOI:** 10.1101/2020.11.06.20227298

**Authors:** Casey Y. Lee, Hany Soliman, Nadia D. Bragagnolo, Arjun Sahgal, Benjamin J. Geraghty, Albert P. Chen, Ruby Endre, William J. Perks, Jay S. Detsky, Eric Leung, Michael Chan, Chris Heyn, Charles H. Cunningham

## Abstract

**Purpose:** Stereotactic radiosurgery (SRS) is used to manage intracranial metastases in a significant fraction of patients. Local progression after SRS can often only be detected with increased volume of enhancement on serial MRI scans which may lag true progression by weeks or months.

**Experimental Design:** Patients with intracranial metastases (N = 11) were scanned using hyperpolarized ^13^C MRI prior to treatment with stereotactic radiosurgery (SRS). The status of each lesion was then recorded at six months post-treatment follow-up (or at the time of death).

**Results:** The positive predictive value of ^13^C-lactate signal, measured pre-treatment, for prediction of progression of intracranial metastases at six months post-treatment with SRS was 0.8 *p* < 0.05, and the AUC from an ROC analysis was 0.77 *p* < 0.05. The distribution of ^13^C-lactate *z*-scores was different for intracranial metastases from different primary cancer types (F = 2.46, *p* = 0.1).

**Conclusions:** Hyperpolarized ^13^C imaging has potential as a method for improving outcomes for patients with intracranial metastases, by identifying patients at high risk of treatment failure with SRS and considering other therapeutic options such as surgery.

## 1 Introduction

Upwards of 20% of cancer patients will develop brain metastases during the course of their illness, many of which become symptomatic [1, 2, 3, 4]. The most common primary tumours responsible for brain metastases include lung cancer, melanoma, renal cancer, breast cancer, and colorectal cancer. The management of brain metastases has become more complex and may involve surgery, stereotactic radiosurgery (SRS), whole brain radiotherapy (WBRT), systemic therapy or best supportive care [5].

Over the past several decades, with improved imaging and radiation therapy technology, stereotactic radiosurgery (SRS) has cemented a role in a significant proportion of patients with brain metastases. SRS has been shown to improve survival in patients with brain metastases over WBRT alone [6] and avoids the the neurocognitive side effects of WBRT [7]. However, local control of brain metastases with SRS decreases with increasing tumour diameter as the dose is dialed back to reduce the risk of radiation injury [8, 9]. Local progression can often only be detected with increased volume of enhancement on serial MRI scans which may lag true progression by weeks or months [10, 11]. Therefore predictive tools for radiation resistance prior to SRS may help identify patients better suited for dose escalation or even upfront surgical resection. If the risk of treatment failure could be better assessed beforehand, some of these patients might be reclassified as candidates for surgery, improving outcomes.

The impact of lactate accumulation on radiosensitivity was only recognized relatively recently [12], but is now well established. Lactate accumulates in malignant tumours through a number of mechanisms, including hypoxia-inducible factor 1 (HIF-1) mediated reprogramming. It has been hypothesized that lactate affects radioresistance by antioxidant properties, inducing angiogenesis, mediating resistance to apoptosis, as well as by stabilizing HIF-1*α* and perpetuating the activation of HIF-1 *independent of* hypoxia [13]. Tumour lactate levels were shown to be inversely correlated with overall and disease-free patient survival in cervical cancer, head and neck squamous cell carcinoma, and glioblastoma in humans [14, 15, 16, 17]. However, these findings were all based on lactate measurements in biopsy or surgical tumour samples, which are not available pre-treatment for intracranial metastases.

Hyperpolarized (HP) ^13^C MRI is an new imaging approach that has the potential to predict radiation treatment failure by probing lactate metabolism. This technique uses a non-radioactive labelled metabolite, [1-^13^C]pyruvate, as a contrast agent. Prior to imaging, the signal of [1-^13^C]pyruvate is amplified by approximately 10,000-fold, or hyperpolarized, via dynamic nuclear polarization (DNP) [18]. While the amplified signal is short-lived (exponential decay constant of *∼* 40s *in vivo*), images of [1-^13^C]pyruvate and its metabolic products such as [1-^13^C]lactate and ^13^C-bicarbonate, can be detected within seconds of injection. Clinical applications of HP ^13^C MRI have been explored in recent years, starting with the first in-human imaging of prostate cancer patients in 2013 [19]. The application of HP ^13^C MRI in patients with brain tumours and metastases is currently an active area of investigation [20, 21, 22].

In this work, patients (N=11) with one or more newly developed intracranial metastases were scanned using hyperpolarized ^13^C MRI prior to treatment with SRS. The mean ^13^C-lactate signal from each lesion was normalized using the consistent pattern of ^13^C-lactate in the brain parenchyma. The status of each lesion was then recorded at the six months post-treatment follow-up (or at the time of death). This enabled estimation of the positive predictive value and receiver operating characteristic (ROC) curve for prediction of lesion progression at six months post-treatment with SRS.

## 2 Materials and Methods

Written informed consent was obtained from patients with intracranial metastases (N = 11) prior to study participation under a protocol approved by the institutional Research Ethics Board and by Health Canada as a Clinical Trial Application. All participants had one or more newly developed intracranial metastasis and were scheduled for SRS immediately following the ^13^C imaging study.

Prior to being positioned head-first in a General Electric (GE) MR750 3.0T MRI scanner (GE Healthcare, Waukesha, WI), a 20-gauge intravenous catheter was inserted into each patient’s forearm. The patient’s head was then secured in the support for the head coil base for a standard 8-channel neurovascular receive array (Invivo Inc.). This support could be docked with either the 8-channel ^1^H array or a home-built single-tuned ^13^C birdcage coil without moving the patient’s head during the study. At the beginning of each exam, localizer images and a reference scan to be used during the image reconstruction [23] were acquired using the scanner’s built-in body coil. The ^13^C birdcage coil was then put in place.

Each subject was injected with a 0.43 mL/kg dose of 250 mM [1-^13^C]pyruvate via an intravenous injection at 4 mL per second using a MEDRAD Spectris solaris injector (Bayer). The doses were prepared within a sterile fluid path and hyperpolarized in a GE SPINLab polarizer equipped with a quality control QC module. The ^13^C image acquisition was initiated upon the completion of a saline flush. All patients tolerated the [1-^13^C]pyruvate injection without any adverse events.

A three-dimensional (3D) spectrally-selective echo-planar imaging (SS-EPI) [24, 23] pulse sequence was used to acquire time-resolved full brain data from [1-^13^C]lactate, [1-^13^C]bicarbonate and [1-^13^C]pyruvate (5-s temporal resolution; 12 time points; 1.5-cm isotropic spatial resolution with a 24 × 24 × 36 cm^3^ field of view).

Following the ^13^C image acquisition, the ^13^C head coil was replaced with an 8-channel ^1^H neurovascular array (Invivo Inc.) for a standard suite of anatomical brain image acquisitions. T_1_-weighted images were acquired using 3D fast spoiled GRE (axial prescription, FOV 25.6 × 25.6 cm^2^, 1-mm isotropic resolution, TR 7.6 ms, TE 2.9 ms, flip angle 11°). Gadolinium enhanced T_1_-weighted images were acquired 2 min after administering a gadolinium dose of 0.1 mmol/kg via hand injection in a subset of patients that required these images for radiation treatment planning. T_2_-weighted images were acquired using T_2_-FLAIR (axial, FOV 22 × 22 cm^2^, in-plane resolution 0.6875 × 0.982 mm^2^, 3-mm slice thickness, TR/TE 8000/120 ms, flip angle 111°).

All ^13^C image reconstruction was performed offline using MATLAB R2018b (The MathWorks Inc., MA, Natick, Massachusetts). Data-driven geometric distortion artifact correction was performed as in [23]. The time-resolved images were summed to compute the area-under-the-curve (AUC) for each metabolite which were stored as final ^13^C images.

The structural parcellation of metabolite signals within the brain was performed using BrainParser [25], which uses T_1_-weighted images to parcellate the brain into the 56 structural regions contained in the LPBA40 atlas. For each subject, the mean ^13^C-lactate signal from each atlas region, *x*_*i*_, was computed and normalized by converting a *z* -score, *z*_*i*_ as in [26]:

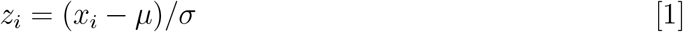

where *µ* and *σ* are the mean and standard deviation of the ^13^C-metabolite signals across the 56 atlas regions for that subject.

A region of interest (ROI) for each metastasis was manually contoured onto each slice of the post-Gd T_1_-weighted images, if available, by a radiation oncologist. In cases where post-Gd T_1_-weighted images were not acquired, manual contouring was performed on the non-contrast T_1_-weighted images or the T_2_-weighted images. The mean ^13^C metabolite signal from each lesion, *x*_*lesion*_, was converted to a *z* -score, *z*_*lesion*_, by using the mean and standard deviation of the background ^13^C-lactate signal in the normal brain, as follows:

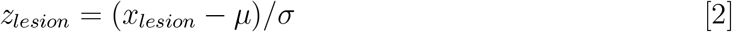

where *µ* and *σ* are as defined above, based on the 56 atlas regions for that patient.

Treated brain metastases were evaluated at the 6 month follow-up or last follow up prior to death. Tumour response was evaluated according to the RANO brain metastases criteria [11]. No cases of radiation necrosis were seen.

## 3 Results

Fig. 1 shows an example ^13^C-lactate and ^13^C-pyruvate image from one of the subjects. The background ^13^C-lactate pattern across cortical regions, expressed as a plot of *z*-scores vs. atlas region, appeared similar to the pattern from healthy participants, but with weaker concordance across patients (Kendall’s *W* = 0.70) as compared with healthy participants (*W* = 0.83) [26]. The pattern is shown for all patients in Fig. 2. Despite the moderate reduction in concordance, the range of lactate *z* -scores from the 56 atlas regions spanned roughly the same range in all subjects, from -3 to 2, which is similar to the range seen in healthy volunteers and sufficiently consistent across patients for the normalization of the tumour ^13^C-lactate signal in this analysis.

**Figure 1:**
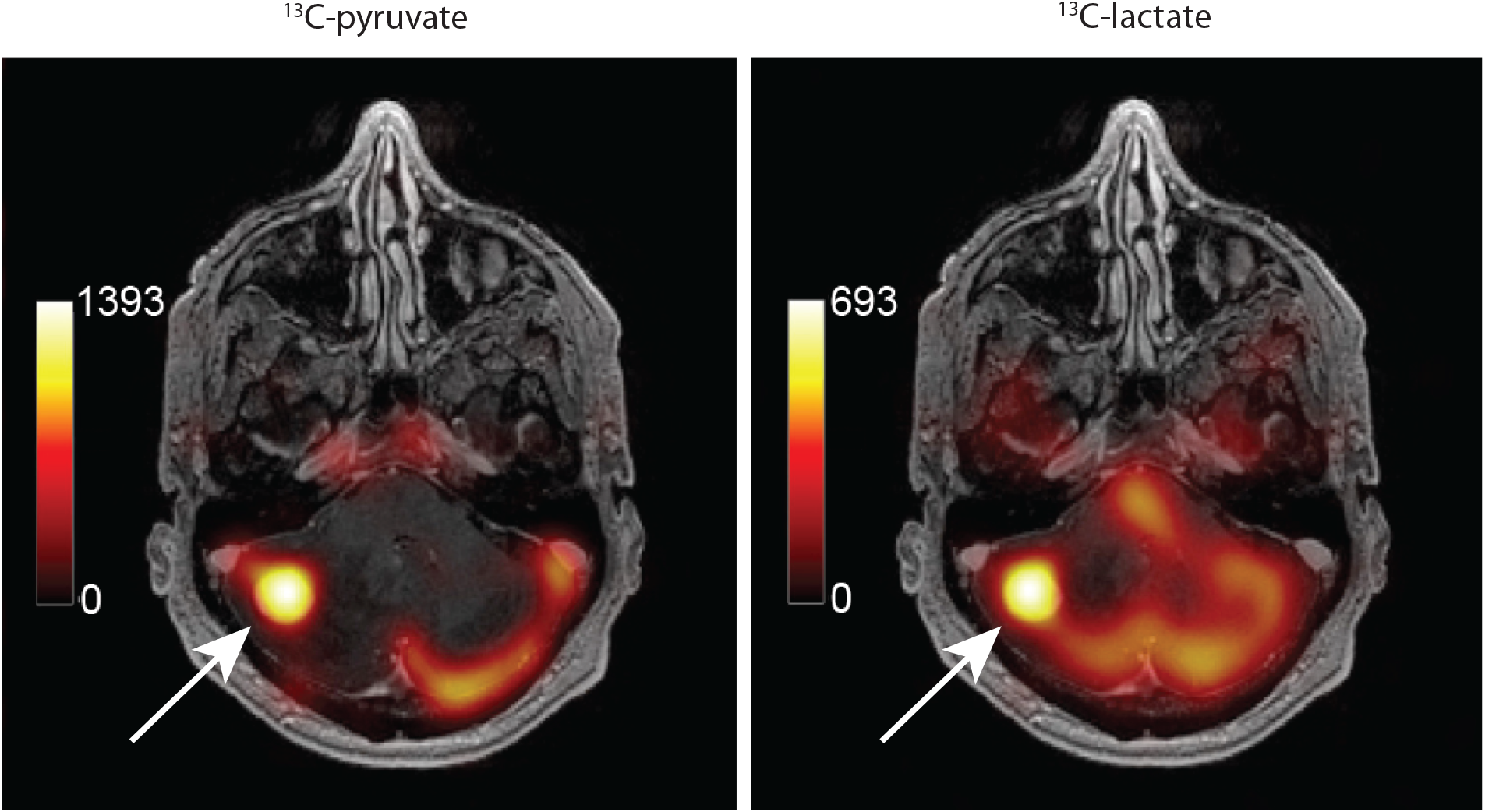
Example ^13^C-metabolite images of an intracranial metastasis (arrows) in a renal cell carinoma (RCC) patient. The metabolite signals are displayed as colour overlays on the corresponding T1-weighted anatomical images in grayscale, and were computed by summing the 12 time-points over the 60 s acquisition window. The treatment of this high-lactate lesion with SRS failed, with local progression prior to the 6-month followup.

**Figure 2:**
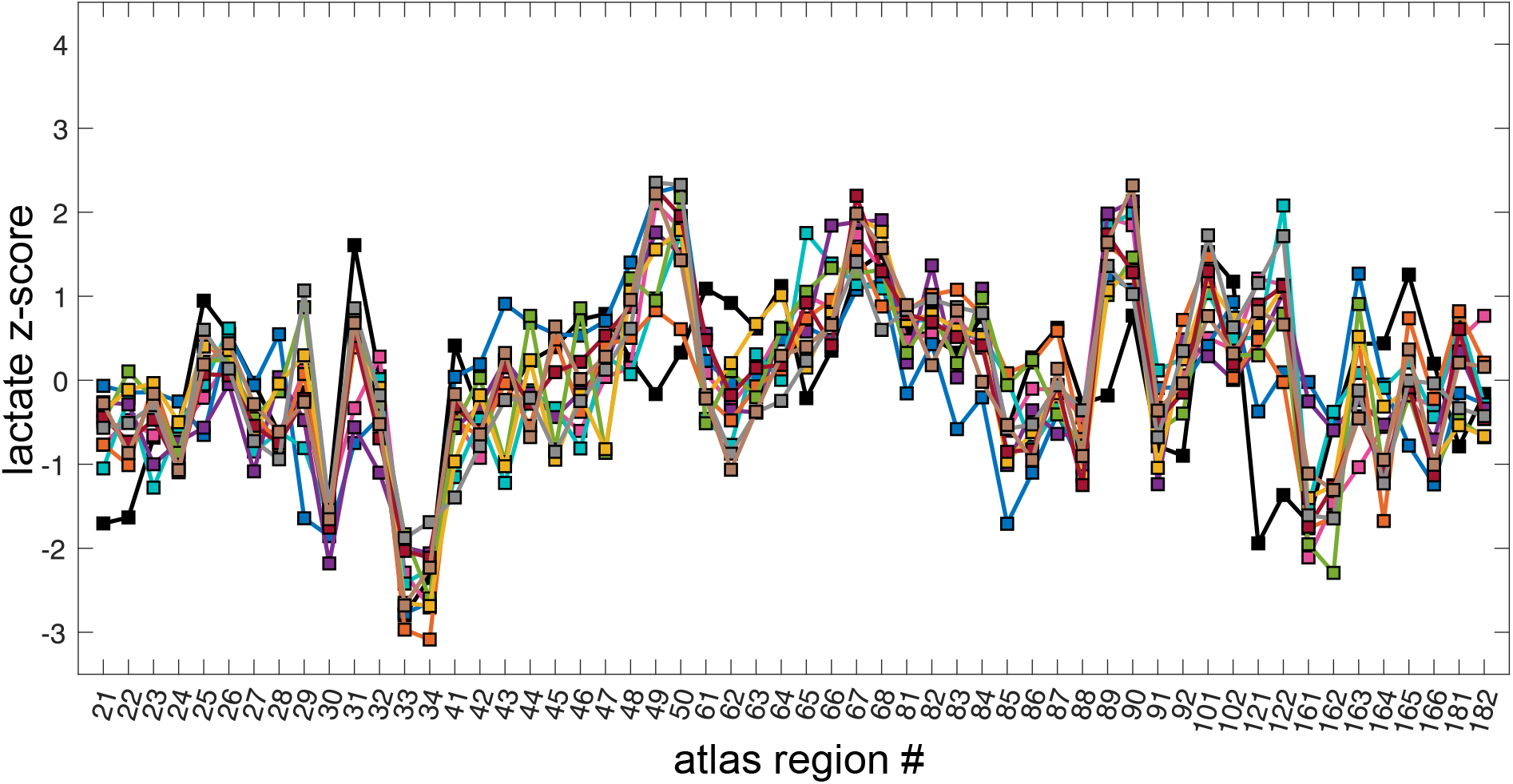
Lactate *z* -scores from 56 LPBA40 atlas regions from 11 patients. The atlas regions are indicated with LPBA40 atlas region numbers. This consistent pattern is used for normalization of the tumour ^13^C-lactate signal.

The metastatic lesions displayed a broad range of lactate *z* -scores, from -2.23 to 3.74, as shown in Fig. 3. A different range of lactate *z*-scores was observed for each of the four primary tumour types: non-small cell lung carcinoma (NSCLC, N=4), renal cell carcinoma (RCC, N=2), breast cancer (N=4) and colorerectal cancer (N=1). To test whether this apparent difference was statistically significant, a 1-way ANOVA was used with the primary cancer type as a categorical variable and the lactate *z*-scores as the dependent variable, giving F = 2.46 and *p* = 0.1.

**Figure 3:**
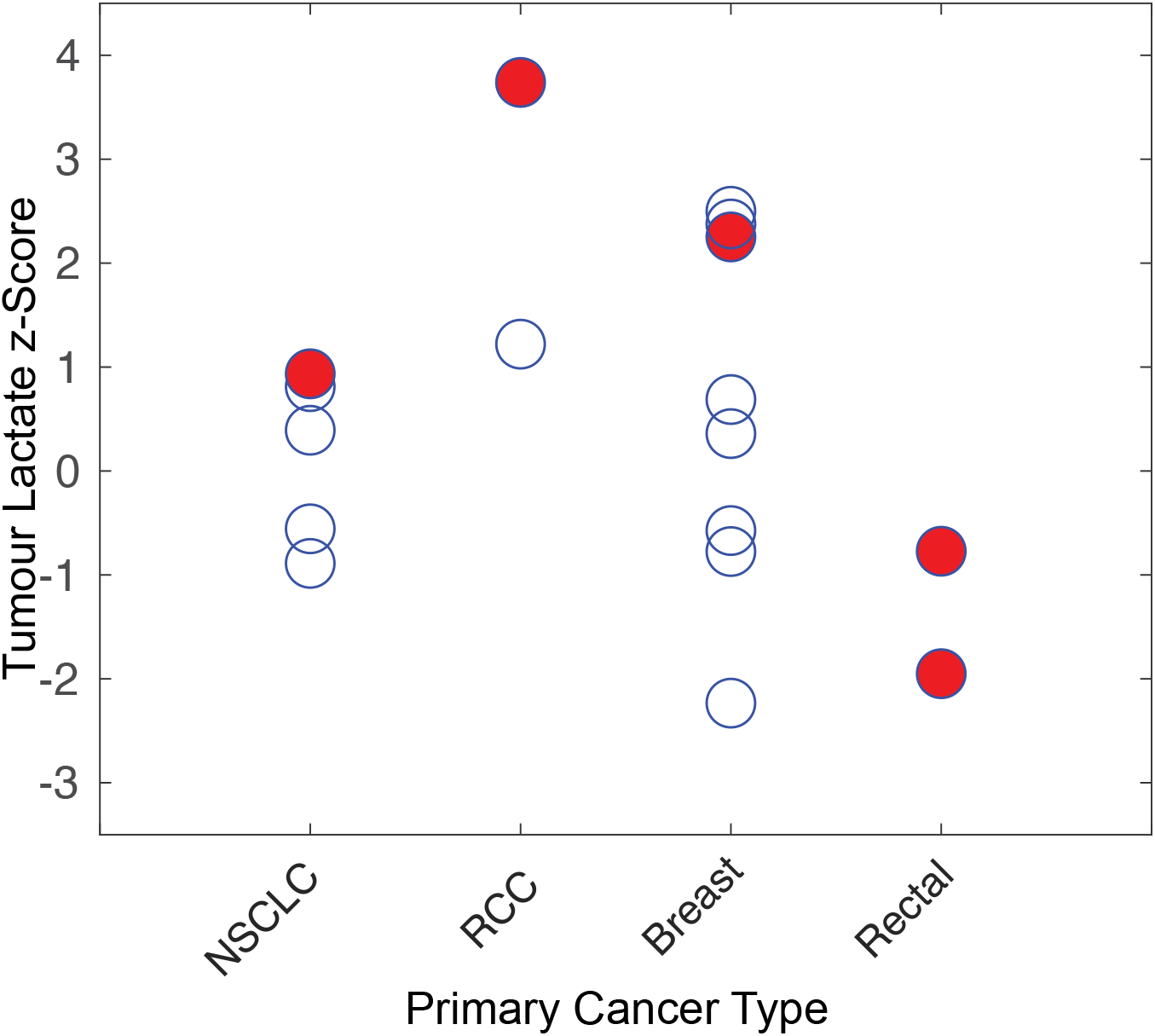
Lactate *z* -scores of newly developed lesions by primary tumour type. The red, solid circles show lesions that progressed, the other open circles are stable/responding lesions. Note the apparent relationship between the higher lactate *z*-scores for each primary cancer type and progression.

Based on this result and the observation that the highest lactate *z*-scores from each primary cancer type appeared to be associated with treatment failure (arrows in Fig. 3), the set of lactate *z*-scores for each primary cancer type was rescaled to a prediction score, ranging from 0 to 1. This allowed the prediction score values from all tumour types to be included in the same ROC analysis, resulting in the ROC curve shown in Fig. 4. The area-under-the-curve (AUC) was 0.77 with an optimal threshold giving a true positive rate of 0.8, a false positive rate of 0.2 and a positive predictive value of 0.8. Based on the equivalency relationship between the AUC and the parameter U derived in the Mann-Whitney U test, as described in [27], the statistical significance of this ROC result was calculated to be *p* < 0.05.

**Figure 4:**
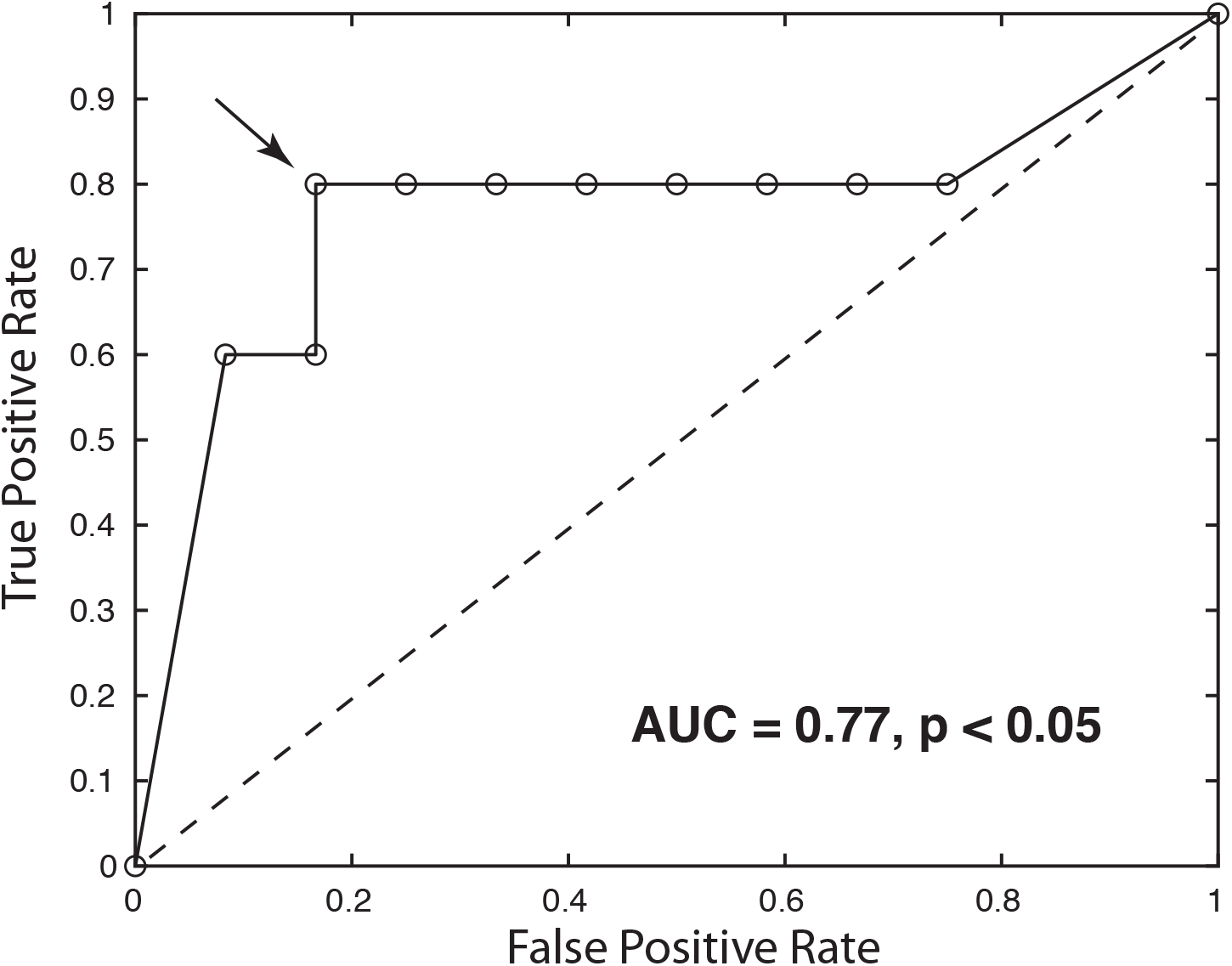
Receiver operating characteristic (ROC) curve for pre-treatment ^13^C-lactate scores as a predictor of treatment failure at 6-months post-treatment with SRS. The optimal threshold is indicated with the arrow, giving a true-positive rate of 0.8, a false-positive rate of 0.2 and positive-predictive value of 0.8 (*p* < 0.05).

## 4 Discussion

The performance of ^13^C-lactate as a predictor of treatment failure was superior to other markers previously investigated for this purpose, such as the apparent diffusion coefficient (ADC) [28] measured *after* SRS. ROC analysis on the prediction of progression using ADC values at 1 week and 1 month post-radiation gave an AUC of 0.704 and 0.748, respectively. Furthermore, a limitation of this study was that lesions from different primary cancers were pooled in the analysis and this required rescaling to equalize the range of values for each primary cancer. In the future, a sufficient number of patients will be scanned so that ROC analysis can be performed on each primary cancer type separately, which will likely improve the AUC.

In this study, a brain parcellation-based approach was used to normalize ^13^C signals from metastatic lesions using the ^13^C signals from other brain regions for reference. This normalization method, based on comparison to the consistent background metabolite pattern in the brain, is a key element of this project as it allows for a ^13^C-lactate measurement of each lesion computed from just the ^13^C-lactate images alone. This is important because elevated ^13^C-lactate signal has been shown to occur alongside elevated ^13^C-pyruvate (substrate) signal under several common conditions such as high tumour vascularity and elevated expression of MCT1 transporters [29]. The conventional normalization methods found in the literature, which involve computation of a lactate-to-pyruvate ratio or fitting a first-order rate constant to time-resolved data, both inherently reduce the number that results from high-lactate/high-pyruvate condition. This condition, in fact, is likely a highly malignant phenotype. One of the lesions that progressed in the preliminary study fell into this category (the highest ^13^C-lactate *z*-score in Fig. 3).

A limitation to this study was the coarse spatial resolution used for the ^13^C imaging (1.5-cm isotropic). This may have lead to contamination of the tumour ^13^C-lactate signal from the surrounding tissues, which would be worse for smaller lesions. Thus the positive predictive value of this method may improve with advances in the data acquisition method that enable higher spatial resolution.

Three lesions across two patients were treated with SRS prior to ^13^C imaging. The lactate *z* -scores from these lesions were -1.13, -1.29 and 1.04. Future studies should investigate whether the observed *z* -scores in previously treated lesions reflect treatment-related metabolic changes or are consistent with the ^13^C-lactate signal levels observed in healthy brain tissues.

## 5 Conclusions

The positive predictive value of ^13^C-lactate signal, measured pre-treatment, for prediction of progression of intracranial metastases at six months post-treatment with SRS was 0.8 *p* < 0.05, and the AUC from an ROC analysis was 0.77 *p* < 0.05. The distribution of ^13^C-lactate *z*-scores was different for intracranial metastases from different primary cancer types (F = 2.46, *p* = 0.1). Hyperpolarized ^13^C imaging has thus shown the potential as a method for improving outcomes for patients with intracranial metastases, by identifying patients at high risk of treatment failure with SRS and considering other therapeutic options such as surgery.

## Data Availability

The data will be made available upon reasonable request to the authors.

## Abbreviations

DE-EPI: dual-echo echo-planar imaging
HP: hyperpolarization
*k*_*pl*_: forward kinetic rates of pyruvate-to-lactate conversion
Lac/Pyr: ratio between intensities of ^13^C-lactate and ^13^C-pyruvate signals
Lac/Bic: ratio between intensities of ^13^C-lactate and ^13^C-bicarbonate signals
NSLSC: non-small cell lung cancer
RCC: renal cell carcinoma
ROC: receiver operating characteristic
*W*: coefficient of Kendall’s concordance

## 6 Author Contributions

CYL, BJG, APC, CHC developed the data acquisition methods, CYL and RE operated the MRI scanner, CYL and CHC performed the image reconstruction and statistical analysis, HS, AS and EL assessed the subjects and performed the ^13^C-pyruvate injections, RE loaded the injector, WJP performed compounding and pharmacy release of the ^13^C-pyruvate doses, CH and HS reviewed the images, CHC and MC designed the study. All authors critically reviewed the manuscript.

## 7 Acknowledgements

The authors thank Julie Green and Sumeet Sachdeva for coordinating the study, and Ruby Endre and Garry Detzler for MR technical support. Funding support from Brain Canada, the Canadian Cancer Society Research Institute and the Canadian Institutes for Health Research grant PJT152928.

---

**Table 1:** (Supplementary) Summary of patients and lesions. Patient ID, primary cancer type, location, lactate *z* -score and clinical status at six months post-treatment followup are listed. Abbreviations: NSCLC, non-small cell lung cancer; RCC, renal cell carcinoma; L, left; R, right.

| Subject ID | Primary Cancer | Location | Lactate z-score | 6-month status |
| --- | --- | --- | --- | --- |
| 11 | NSCLC | brainstem (pons) | -0.89 | stable |
| 12 | RCC | R cerebellum | 3.74 | progressed |
| 20 | NSCLC | L occipital lobe | 0.39 | responding |
| 24 | Breast | R parietal | 2.50 | responding |
| 24 | Breast | R temporal | -0.78 | stable |
| 24 | Breast | L frontal | -1.13 | pre-treated |
| 27 | NSCLC | R frontal | 0.81 | responding |
| 27 | NSCLC | L frontal | -0.56 | responding |
| 34 | NSCLC | L cerebellum | 0.94 | progressed |
| 36 | Rectal | L cerebellum | -0.77 | progressed |
| 36 | Rectral | R cerebellum | -1.95 | progressed |
| 37 | RCC | L cerebellum | 1.22 | stable |
| 37 | RCC | R cerebellum | -1.29 | pre-treated |
| 37 | RCC | L occipital | 1.04 | pre-treated |
| 39 | Breast | R occipital | 0.69 | responding |
| 39 | Breast | M cerebellum | 2.25 | progressed |
| 40 | Breast | L occipital | -2.23 | responding |
| 47 | Breast | R occipital | 2.38 | responding |
| 47 | Breast | R frontal | -0.57 | responding |
| 47 | Breast | L frontal | 0.36 | responding |

